# A Claims-Based Machine Learning Classifier of Modified Rankin Scale in Acute Ischemic Stroke

**DOI:** 10.1101/2025.02.06.25321827

**Authors:** Mamoon Habib, Rafaella Cazé de Medeiros, Syed Muhammad Ahsan, Aidan McDonald Wojciechowski, Maria A. Donahue, Deborah Blacker, Joseph P. Newhouse, Lee H. Schwamm, M. Brandon Westover, Lidia MVR Moura

## Abstract

**Background:** We developed a classifier to infer acute ischemic stroke (AIS) severity from Medicare claims using the Modified Rankin Scale (mRS) at discharge. The classifier can be utilized to improve stroke outcomes research and support the development of national surveillance tools.

**Methods:** This was a multistate study included all participating centers in the Paul Coverdell National Acute Stroke Program (PCNASP) database from nine U.S. states. PCNASP was linked to Medicare data sets for patients hospitalized with AIS, employing demographics, admission details, and diagnosis codes to create unique patient matches. We included Medicare beneficiaries aged 65 and older who were hospitalized for an initial AIS from January 2018 to December 2020. Using Lasso-penalized logistic regression, we developed and validated a binary classifier for mRS outcomes and as a secondary analysis we used ordinal regression to model the full mRS scale. Performance was evaluated on held-out test data using ROC AUC, ROC Precision-Recall, sensitivity, and specificity.

**Results:** We analyzed data from 68,636 eligible patients. The mean age was 79.5 years old. 77.5% of beneficiaries were White, 14% were Black, 2.6% were Asian, and 2% were Hispanic. The classifier achieved an ROC AUC score of 0.85 (95%CI: 0.85-0.86), sensitivity of 0.81 (95%CI: 0.80-0.81), specificity of 0.73 (0.72 - 0.74), and Precision-Recall AUC of 0.90 (95%CI: 0.90-0.91) on the test set.

**Conclusion:** Among Medicare beneficiaries hospitalized for AIS, the claims-based classifier demonstrated excellent performance in ROC AUC, Precision-Recall AUC, sensitivity, and acceptable specificity for mRS classification.

**Clinical Perspective:** *What Is New?:*

- Developed a novel claims-based classifier to infer acute ischemic stroke (AIS) severity using the Modified Rankin Scale (mRS) at discharge.
- Integrated Medicare claims with clinical data from the stroke registry, utilizing penalized logistic regression for both binary and ordinal classification.

*What Are the Clinical Implications?:*

- Provides a robust tool for assessing stroke severity, which can enhance stroke outcomes research and quality improvement initiatives.
- Supports the development of national surveillance tools, potentially guiding clinical decision-making and resource allocation in stroke care.

**Research Perspective:** *What New Question Does This Study Raise?:*

- How can claims-based severity classifiers be effectively integrated into existing stroke research and clinical practice to enhance outcome measurement?
- To what extent is the classifier generalizable to diverse populations beyond Medicare beneficiaries?

*What Question Should be Addressed Next?:*

- Future research should evaluate the impact of incorporating such classifiers into risk adjustment processes and their effect on long-term stroke outcomes.
- Investigate whether similar modeling approaches can be adapted for other patient groups and healthcare settings to improve surveillance and treatment strategies.

## INTRODUCTION

Every 40 seconds, someone in the United States (U.S.) has a stroke.^1^ Stroke is one of the leading causes of long-term disability, affecting about 795,000 people in the U.S. annually.^2^ Acute ischemic stroke (AIS) severity can be variable, with a significant portion of discharged patients presenting with declining functionality, leading to increased needs for rehabilitation and admission to nursing facilities.^3^ Both modifiable (i.e., obesity, diabetes, cardiovascular disease, certain medications, physical inactivity, etc.) and non-modifiable stroke risk factors (i.e., age, sex, race/ethnicity, genetics) can help determine prognosis, which is crucial for early tailored intervention.^4^

Functional outcome prediction in AIS impacts the quality of patient care decisions.^5,6^ Recent advances in computational and software technologies have greatly impacted the rise of Machine learning (ML) studies, offering more precise outcome measures.^7–9^ ML models have identified several crucial factors to predict and classify functional outcomes, such as an initial National Institutes of Health Stroke Scale (NIHSS) score, age, fasting blood glucose, creatinine levels, and the modified Rankin Scale (mRS).^10,11^ mRS has been widely used to assess AIS severity and clinical prognosis in electronic health records (EHRs) and registries.^12^ The creation of models and classifiers can be personalized to assess outcomes in AIS patients, including the classification of mRS.^8,9,13^ However, limited valid measures of stroke severity have hindered national, large-scale, claims-based studies.^14^

Despite this limitation, claims data may offer indirect clues about a patient’s level of disability based on the types of claims filed. Leveraging a dataset that links claims to mRS scores, we explored whether supervised ML could develop a classifier to infer mRS from claims information. Such a model could enable the personalization of outcome assessments for AIS patients and the classification of mRS in large, claims-based studies, thereby configuring a tool for national surveillance of stroke severity.

We linked the Paul Coverdell National Acute Stroke Program (PCNASP) and Medicare claims-based inpatient data of older adults presenting with AIS to develop and validate the mRs classifier of stroke severity at discharge.

## METHODS

The Medicare data supporting this study’s findings are collected routinely by The Centers for Medicare & Medicaid Services (CMS) for billing purposes and were made available by CMS with no direct identifiers. All results were aggregated following CMS Cell Suppression Policies. Restrictions apply to the availability of these data, which were used under license for this study. Medicare data are available through CMS with their permission. PCNASP data are available through the CDC with their permission.

This study was approved by the Mass General Brigham Institutional Review Board’s (IRB) ethical guidelines and followed the Strengthening the Reporting of Observational Studies in Epidemiology (STROBE) guidelines for observational studies^15^ and the transparent reporting of multivariable prediction models developed or validated using clustered data (TRIPOD)^16^ and the updated guidance for reporting clinical prediction models that use regression or machine learning methods (TRIPOD-AI).^17^

### Study Design

We conducted a retrospective analysis of claims data from AIS patients using a sample from nine large U.S states. We aimed to develop and validate a classifier based on claims data that infers mRS at discharge.

### Data Source

We accessed data from the PCNASP registry and Medicare Claims data. PCNASP collects data on stroke cases and captures discharge mRS scores reported by clinicians or hospital staff.^18^ The PCNASP registry includes information from 2008 to 2020from the following. states: California; Georgia; Massachusetts; Michigan; Minnesota; New York; Ohio; Washington; and Wisconsin.

We then matched the PCNASP data on individuals aged 65 or older with data from fee-for-service Medicare, a national health insurance program administered by the Centers for Medicare & Medicaid Services (CMS).^19^ The Medicare Provider Analysis and Review (MEDPAR) files contain extensive information about these beneficiaries, including patient demographics, admission and discharge dates, diagnosis, procedure codes, provider identifiers, and comorbidities.^20^

### Study Population

We analyzed Medicare claims data for beneficiaries aged 65 and older hospitalized for AIS from January 2018 to December 2020. We included beneficiaries who were enrolled in traditional Medicare Part A (inpatient hospital insurance; care in a skilled nursing facility, hospice care, and some home health care) and Part B (physician and other medical provider services; outpatient care, medical supplies, and preventive services) who had mRS values documented in the PCNASP clinical database (based on ICD-10 code information).

We used a multi-step exclusion and inclusion process to refine our patient population. First, we excluded patients with missing mRS scores and deceased patients in the PCNASP data and then linked the remaining data with Medicare claims data. We found patients with a diagnosis of AIS in the Medicare claims data during 2018-2020 and used only their first stroke encounter. We next created two groups based on the availability of an mRS score for any stroke (Supplemental Figure 1). The first group included patients admitted to the hospital with a ≥90% or more completion rate of mRS, while the second group included patients admitted to hospitals with less than <90% of mRS completion. We used 20% of the first group and all of the second group as a training sample; the remaining 80% of the first group was set aside as an independent test sample.

### Linking Databases

Because there were no unique patient identifiers common to both databases, we applied a matching strategy to link individuals in the PCNASP and Medicare datasets.^21^ For this linkage we used variables such as age, gender, admission and discharge dates, diagnosis code, hospitals, and state. After linkage, we retained patients with unique matches, excluding cases where PCNASP IDs corresponded to multiple Medicare Beneficiary IDs and vice versa. Due to limited access to baseline institutionalized (non-outpatient) data, we excluded patients transferred from another hospital, skilled nursing facility (SNF), or other healthcare facilities.

### Variables

We included demographic variables, medical history, treatments, and discharge outcomes. Most variables were extracted from the MEDPAR files. Those not included in MEDPAR were extracted from hospital level data by linking MEDPAR data with provider-level data and included variables such as bed size and hospital location, category and level. We included two stroke-related variables for inpatient conditions and procedures such as tissue plasminogen activator (tPA) and endovascular treatment. We used the value “1” if the condition or procedure was present and the value “0” if not. For continuous variables such as age and length of stay, we standardized their values. Categorical variables, such as race and admission type, were converted into dummy variables for use in the model. We used the variables included in the Chronic Conditions Warehouse (CCW) algorithms from Medicare to determine comorbidities and relevant patient medical history in our patient population.^22^ CCW flagged 27 chronic conditions for each beneficiary within the study period, which we used to determine if the beneficiary had any comorbidities. We selected the first-ever criteria a beneficiary met for the chronic condition.

### Construct of Interest (Endpoint)

Our primary endpoint was the accurate classification of mRS at discharge. We dichotomized the mRS scale into “favorable” if valued as equal or less than 2 (from no symptoms to slight disabilities) and “unfavorable” if the mRS score was > 2 (interval from moderate disability to death).^12,23^

As a secondary analysis, we developed ordinal classifiers using the previous sampling approach to obtain more granularity among mRS categories. The two approaches of ordinal classification consist of a full mRS scale, one represented by 0: no symptoms; 1: no significant disabilities, despite symptoms; 2: slight disabilities; 3: moderate disability; 4: moderate to severe disability; 5: severe disability; 6: death.^23, 24^ The second ordinal model consists of the same full scale but excludes the death category.

### Model Development

#### Primary analysis - Binary Classifier

The binary classifier outputs probabilities for each class. A threshold of 0.5 was used to convert the probabilities into binary values. Predictions with a probability greater than or equal to 0.5 were assigned to the unfavorable mRS category, and those below 0.5 to the favorable class.

For development of our binary classifier, binary logistic regression with a lasso penalty was trained to predict the binary mRS category (favorable vs unfavorable). The best hyperparameters were determined through a grid-search hyperparameter tuning process. The hyperparameters included a range of the inverse regularization strength C (10⁻ ⁴ to 100), tolerance values (1e-4 to 1e-1), maximum iterations (5000 to 50000), solver methods (’liblinear’ and ’saga’), and class weight settings (None and Balanced). The hyperparameters that generated the largest area under the receiver operator characteristic curve (ROC AUC) were chosen. Stratified 5-fold cross-validation was used to evaluate the classifier’s performance within the training set. The model was separately evaluated on the test set, which was not used in model development.

#### Secondary analysis - Ordinal Classifier

We also trained a classifier on the full-scale mRS values using ordinal regression. The ordinal regression model outputs probabilities for each class. To assign class labels, we selected the class with the maximum predicted probability.

We fitted the model as a parallel classifier with a logit link and Lasso L1 penalty using the ordinalNet R package. Grid-search hyperparameter tuning was performed on the training dataset to select the best model based on lambda and family values. We defined a sequence of lambda values (ranging from 0.001 to 0.01) and multiple family values (cumulative, acat, sratio, cratio).

For each family type in the classifier, models were fitted across a range of lambda values and log-likelihood was used to evaluate model performance. The optimal lambda for each family type was selected as the value that achieved the highest log-likelihood, once we selected the optimal family type and lambda value, we refitted the final classifier on the training data with the chosen parameters. We tested the refitted model on the test dataset to check for its generalizability.

### Performance Metrics

For both primary and secondary analyses, we evaluated classifier’s performance using ROC AUC and Area Under the Precision-Recall Curve (PR AUC) to assess the model’s ability to distinguish between classes. Sensitivity and specificity, were included to evaluate the model’s ability in identifying true positives and true negatives.

To calculate confidence intervals (CI) for our performance metrics, we performed 10,000 iterations of bootstrap random sampling with replacement in each iteration. We created a distribution for each metric and calculated 95% confidence intervals to show the classifier’s performance variability.

## RESULTS

### Characteristics of the samples

We assessed 295,241 hospital admissions for AIS between January 2018 and December 2020 for eligibility. After applying our inclusion and exclusion criteria, our sample included 68,636 unique Medicare beneficiaries who were 65 years old or older with a first admission for AIS and available discharge mRS scores. We obtained distinctive patient hospital encounters with < or ≥ 90% completion of the mRS (N= 33,654 and N= 34,982, respectively) (Supplemental Figure 1).

The mean age for the full sample was 79.53 (SD 8.7), and 77.5% of beneficiaries were White, 14% were Black or African American, 2.7% were Asian, and 2% were Hispanic (Table 1). The mean age for our test data was 79.76 (SD 8.7). Approximately 91% of our patient sample was admitted through emergency care. Regarding discharge disposition, the test set data was more evenly distributed between home, SNFs, and inpatient rehabilitation facilities with 28%, 23%, and 19%, respectively, followed by interventions, such as receipt of tissue plasminogen activator and endovascular intervention. The remaining percentage was distributed between approximately 100 other discharge disposition variables. Concerning comorbidities, 71% of beneficiaries had hypertension, 39% diabetes, and 29% congestive heart failure. A further breakdown of the full sample, training, and test set demographics can be found in Table 1. We used 63 covariates to predict a scale score, such as demographics, medical history, treatments, and discharge outcomes (a list can be found in Figure 1 and Supplement Table 5).

**Table 1.**
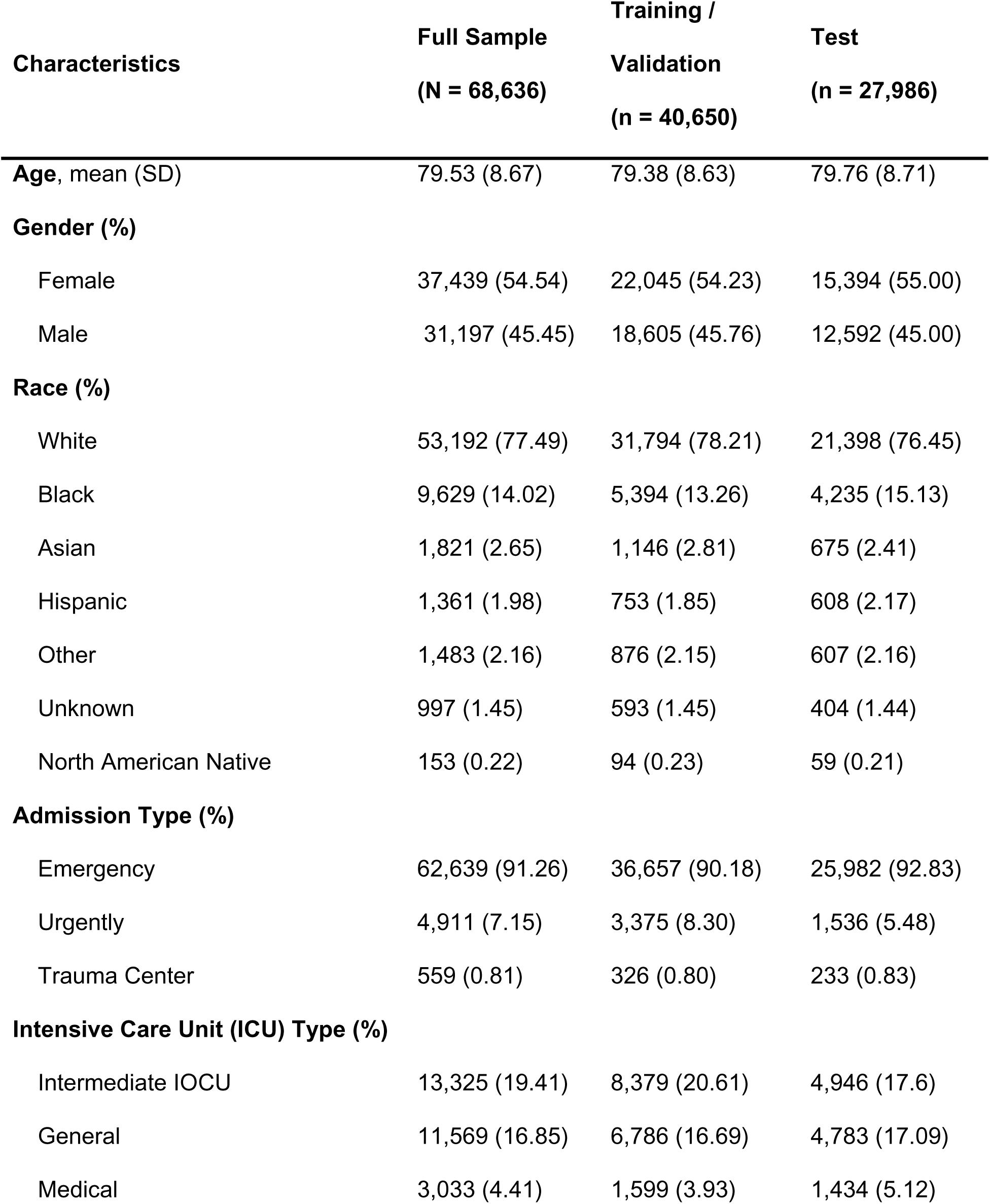
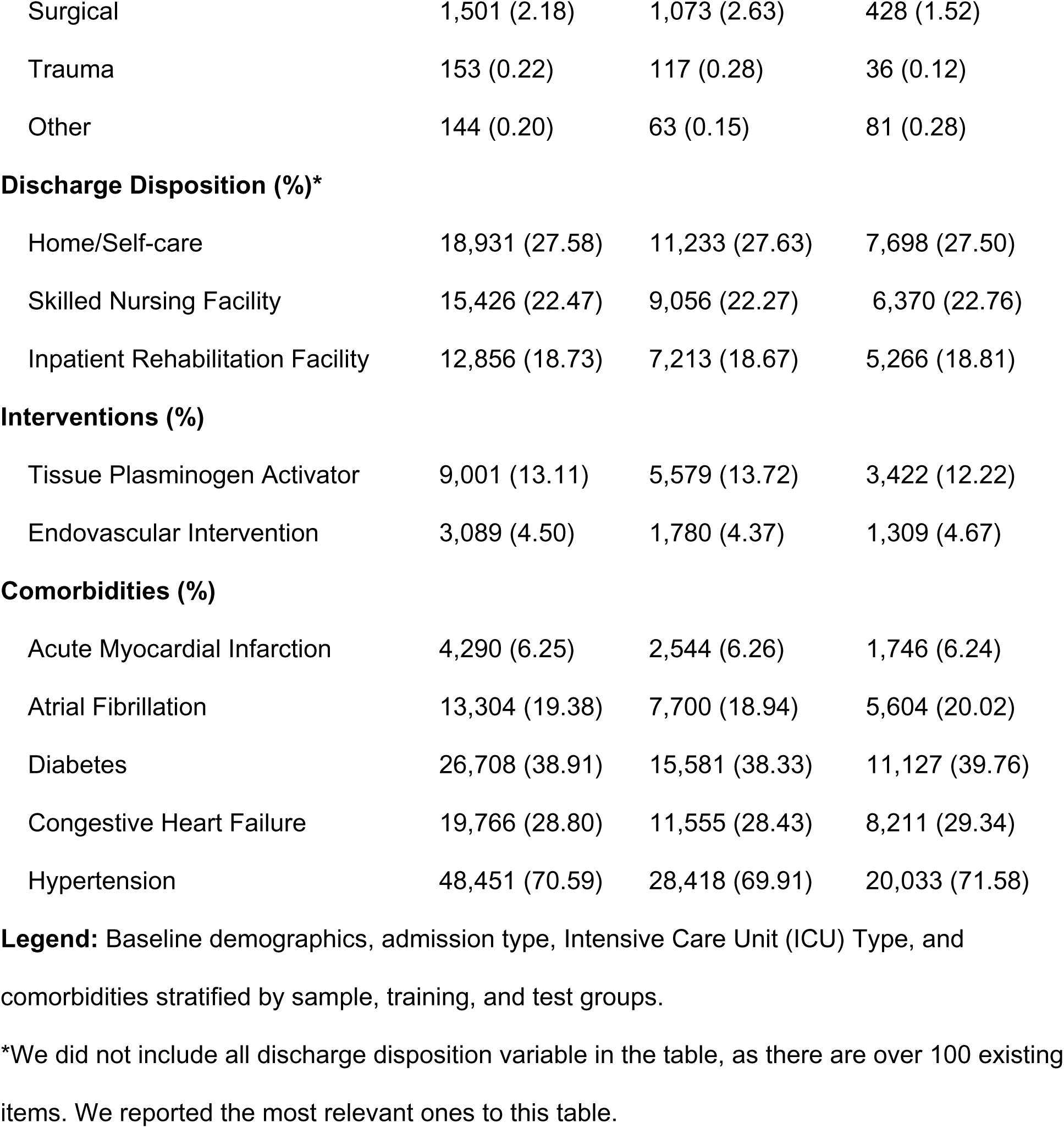
Demographic Characteristics. Baseline demographics, admission type, Intensive Care Unit (ICU) Type, and comorbidities stratified by sample, training, and test groups. *We did not include all discharge disposition variable in the table, as there are over 100 existing items. We reported the most relevant ones to this table.

**Figure 1.**
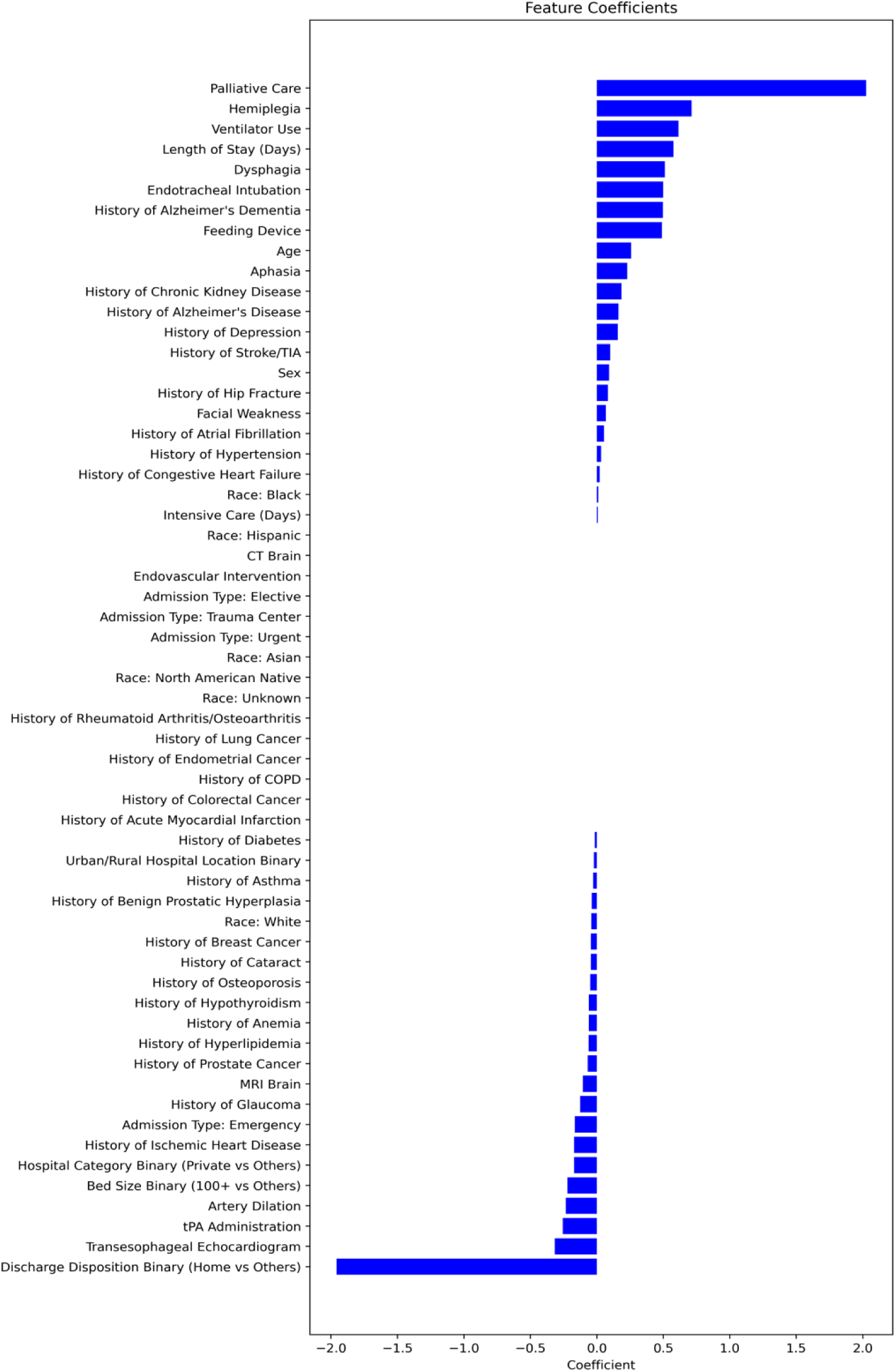
Model Features. The full list of the classifier’s features and their coefficient values. COPD: Chronic obstructive pulmonary disease; ICU: Intensive Care Unit; tPA, tissue plasminogen activator; CT, computed tomography; MRI, Magnetic resonance imaging.

### Binary Classifier

On the held-out test data, our binary classifier achieved an ROC AUC score of 0.85 (95%CI: 0.85 – 0.86, Figure 2), sensitivity of 0.81 (95%CI: 0.80 – 0.81), specificity of 0.73 (0.72 - 0.74), and Precision-Recall AUC of 0.90 (95%CI: 0.90 – 0.91, Figure 3). Figure 1 shows the model’s feature coefficients sorted/ranked by their contribution to its predictions. Palliative care was the strongest predictor (2.02) of unfavorable mRS outcomes. Similarly, coded hemiplegia (0.71), and the use of ventilator during the AIS hospitalization (0.61) were strong predictors of unfavorable outcomes. Several features were also associated with a lower likelihood of unfavorable outcomes. For instance, binary discharge disposition (home vs others) had the strongest negative coefficient (-1.95), suggesting that favorable discharge outcomes strongly predict better recovery. Transesophageal echocardiogram (-0.31), and tPA administration (- 0.25), were associated with favorable outcomes.

**Figure 2.**
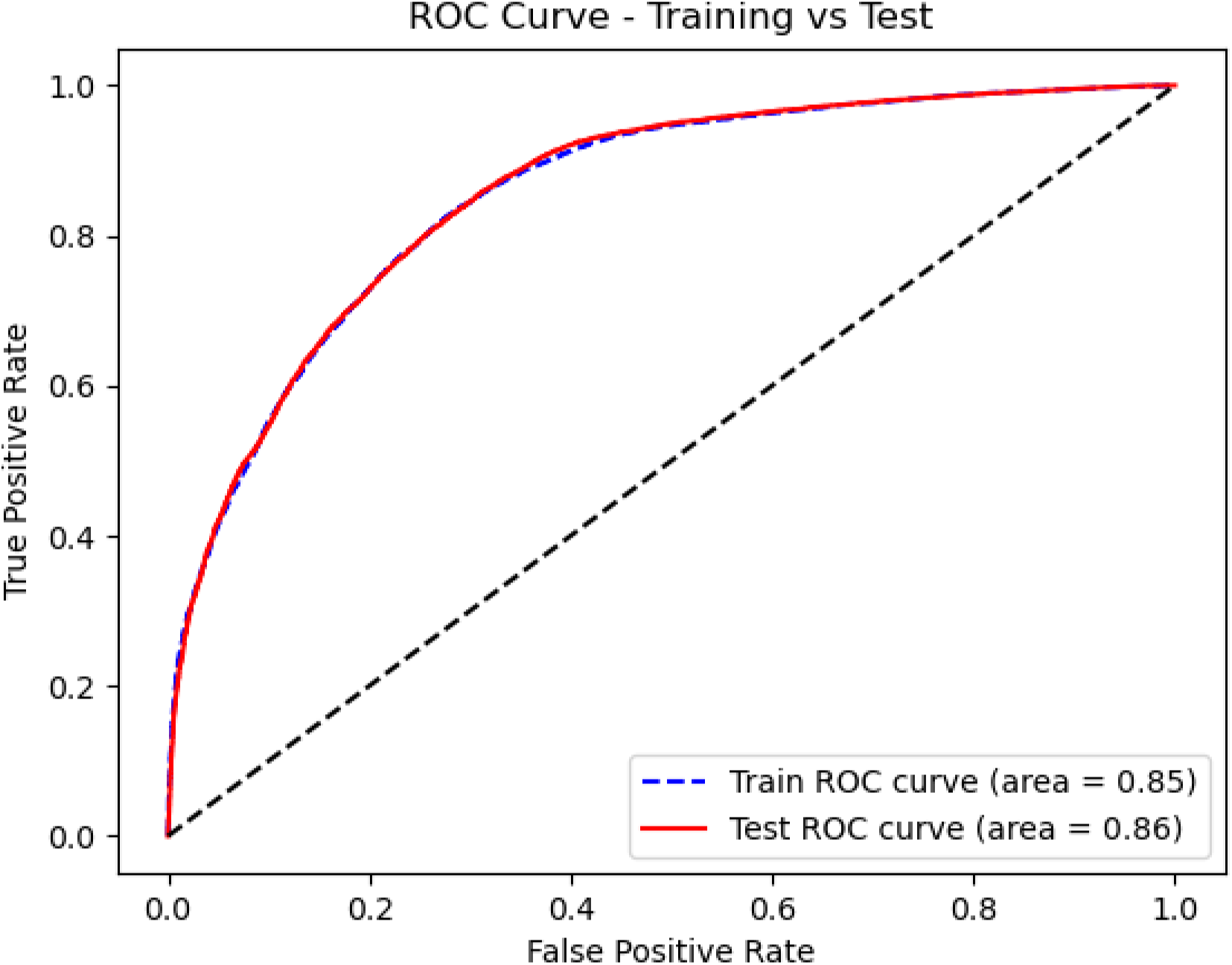
ROC (Receiver Operating Characteristic) Curve. Comparison of the ROC in both the training and test sets of the classifier.

**Figure 3.**
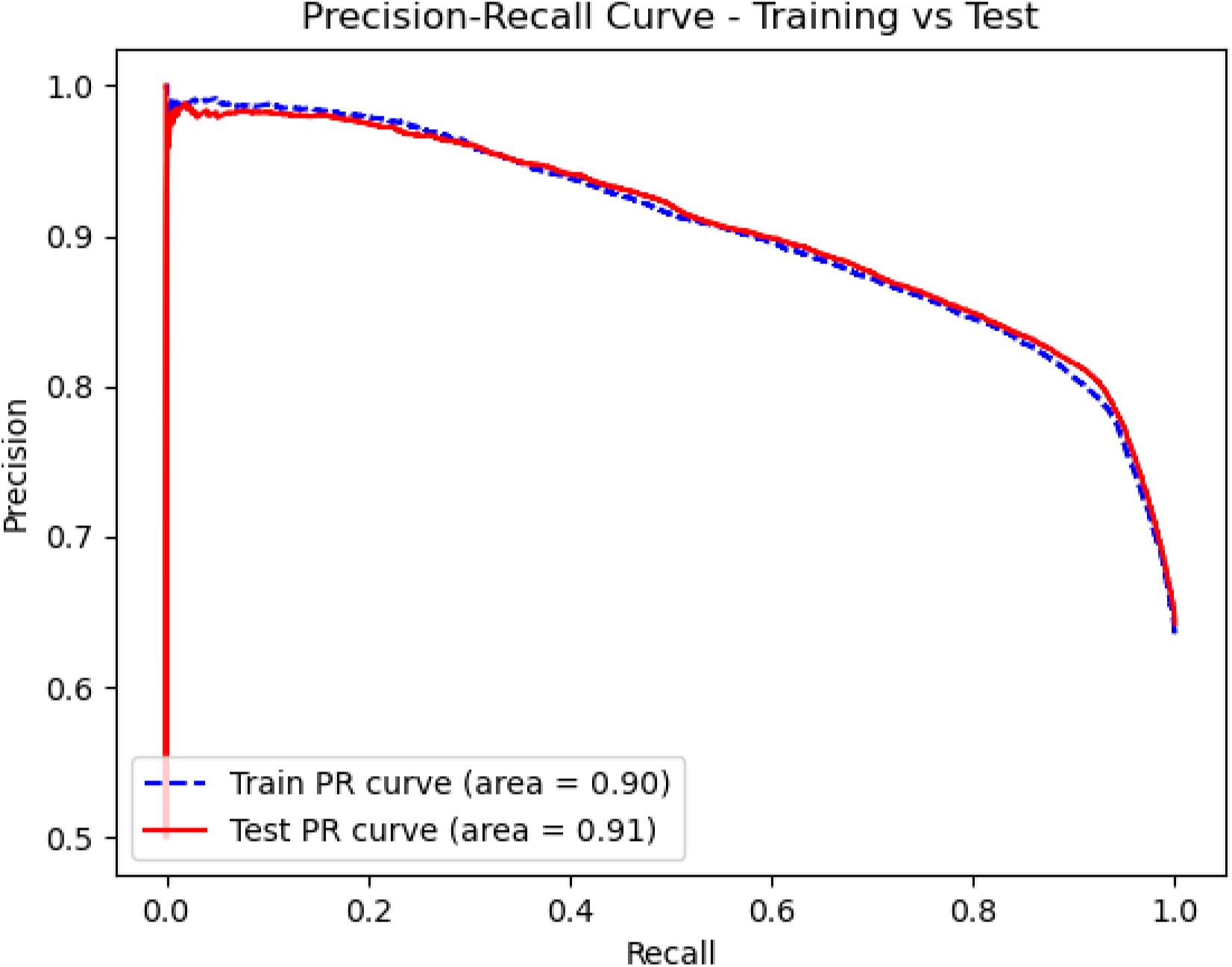
Precision-Recall Area Under the Curve for Binary Classifier. Comparison of Precision-Recall Curve of the classifier in the training and test sets.

### Ordinal Classifier

For our secondary analysis, the ordinal model’s overall performance on the test data is presented in Table 2. The model demonstrates a stronger ability to distinguish between mRS scores 0 (No Symptoms) and 5/6 (Severe Disability/Death) compared to its performance in differentiating intermediate outcomes (1–4) [see Supplementary Figure 2].

**Table 2.** Full-Scale Ordinal Model Performance. Performance Metrics from Full-Scale Ordinal. We report micro-average ROC AUC and Precision-Recall AUC.

| Metric | Score [CI] |
| --- | --- |
| ROC AUC | 0.81 [0.80 – 0.81] |
| Precision-Recall AUC | 0.39 [0.37 – 0.39] |
| Sensitivity | 0.42 [0.41-0.2] |
| Specificity | 0.89 [0.88 - 0.89] |

Classes 2 (Slight Disability) and 3 (Moderate Disability) showed the lowest ROC AUC and PR AUC scores. Supplementary Figure 4 presents a box plot of grouped probabilities, highlighting how the model conflates mRS scores 2 and 3 with mRS score 4. The model’s ability to distinguish between mRS scores 0 (No Symptoms) and 5/6 (Severe Disability/Death) is higher compared to its performance in differentiating intermediate outcomes (1–4) [see Supplementary Figure 2].

Additionally, we excluded death to evaluate whether the model’s performance improves in predicting intermediate outcomes 2 and 3, however, no significant changes in performance were observed. The model’s performance is presented in the supplementary section. The coefficients from both ordinal models (see Supplementary Tables 6 and 7) were consistent with those observed in the binary model. For instance, in the full-scale mRS ordinal model, discharge disposition [i.e., discharged home] (coefficient = 1.99) increased the odds of falling into a lower (better) mRS category, whereas palliative care (coefficient = –2.72) increased the odds of a higher (worse) category.

## DISCUSSION

Considering the clinical burden of AIS and its influence on patient mortality, rate of disability, medical complications, and healthcare expenditures, it is fundamental to monitor the impact, severity, and prognosis of this condition.^1,31,34^ Our interpretation of the identified factors driving the classification highlights their strong face validity and consistency with existing literature as they align with clinical expectations and prior studies. Palliative care, hemiplegia, endotracheal intubation, and feeding device usage were strong predictors of unfavorable mRS outcomes, which is consistent with established knowledge on poor prognostic factors in acute ischemic stroke. Similarly, favorable discharge disposition (e.g., discharged home), tPA administration and brain imaging (CT or MRI) were associated with better outcomes, reinforcing the importance of early and effective stroke management.

We developed and validated a claims-based classifier to accurately identify stroke severity measured by mRS at discharge in patients aged 65 or older who experienced AIS. By leveraging administrative claims data, our classifier demonstrates strong predictive performance, achieving excellent accuracy for categorizing stroke severity. This tool holds significant potential for facilitating large-scale research on stroke outcomes and improving national surveillance efforts, enabling more effective monitoring of stroke care quality and recovery outcomes.

Validated claims-based classifiers for AIS surveillance are also important for observing geographic trends and are essential for population health research, which in turn can inform public health policy and national guidelines to improve clinical practice.^3^

Previous studies have utilized ML methods for stroke functional outcome assessment.^5,13,28^ Joon Nyung Heo et al. measured mRS 90 days after hospital discharge using three learning algorithm models: deep neural network, random forest, and logistic regression.

The study had similar results with the logistic regression model (AUC 0.85), while the best performance was by the deep neural network model (AUC 0.88) ^28^ In our study, logistic regression for mRS classification at discharge yielded positive results with the ROC AUC score of 0.85, reiterating the results seen in other models.^5,13,28,31^

Most importantly, the previous studies were limited by selection bias due to their sampling from single regions of the US.^5,13,28,31^ Our study overcomes this challenge by including a national, large-scale sample with representation of patients and practices from nine U.S. states spanning all regions of the US. Therefore, our cohort provides a more robust, inclusive, and representative claims-based classifier for beneficiaries with AIS than has been heretofore available.

Prior studies creating mRS stroke-severity classifiers used a random assignment approach within hospitals to create training and test sets.^9,13^ This approach is potentially biased because random sampling does not account for hospital-level patterns in patient intake and reporting. We addressed this by categorizing the training and test data sets depending on whether hospitals reported < or ≥ 90% mRS completion. We only used data from those with ≥90% mRS completion as the test set, with a random 20% allocated to the training set for representativeness, allowing the classifier to be trained and tested with higher-quality data and partially accounting for potential bias in random sampling.

Furthermore, our study used binary and ordinal regression methods to classify the mRS score in AIS patients. Binary analyses yield results that are more easily interpreted by examining the absolute risk reduction between the two groups but do not exploit the within-group variation .^23^ We therefore also implemented an ordinal approach to achieve better use of the dataset.^23,32^ The use of the ordinal method increased statistical power and decreased loss of information when compared with previous studies.^5,33^

Other research groups have focused on validating admission stroke severity, such as electronic health record (EHR)-based classifiers of NIHSS at admission.^25^ This is important work, as classifiers of stroke severity at admission can inform resource allocation while patients are admitted and guide other care measures. However, we focused on leveraging claims data to classify stroke functional outcomes at discharge using the mRS. The mRS is important because it provides information on patient functional outcomes, which can inform the prioritization of post-discharge stroke care allocation and predictions of long-term outcomes, among other applications.^26,27^ The score’s ability to predict the level of functionality makes it an essential tool for national-level surveillance using administrative databases.^5^

### Limitations

While we used a nationally representative stroke registry covering nine U.S. states and its major stroke centers linked to administrative claims data, results may not be generalized to states not included in our data set or smaller community healthcare centers. In addition, our selection of older adults ≥65 covered by fee-for-service Medicare may not represent other patient populations. Slightly over half of eligible Medicare beneficiaries are now enrolled in Medicare Advantage “Part C” instead of traditional Medicare. Beneficiaries must also be enrolled in Parts A and B, as well as Part B premium. Recent studies have shown that enrollment in lower-cost Medicare Advantage plans has increased among low-income and racial/ethnic minorities.^35^ Future studies assessing these groups would benefit these populations.

We excluded also 12,894 patients transferred from another hospital, skilled nursing facility (SNF), or other healthcare facilities from the analytical sample, which may have omitted a subset of the AIS population with a higher burden of baseline comorbidities. We selected this approach due to limited access to predictor data from these groups. Including these patients could have enhanced classifier representativeness and performance by increasing the sample size and introducing greater variability. Nevertheless, our classifier demonstrated high performance while capturing a broad and still nationally representative segment of the AIS population.

We were limited by data availability for the Medicare and PCNASP datasets. While utilization of administrative claims linked to data registries represents a vast source of information for research purposes,^36^ some inherent limitations (e.g., human-type errors of scores and clinical scales and missing data e.g., missing mRS scores and other stroke-related variables) are surely present. Despite these limitations, national administrative claims data remains valuable in representing large-sized populations and their reflections.^37,38^

Lastly, the replicability of our classifier can present some challenges, for example, requiring at least two databases to perform linkage of common unique identifiers and extract multiple variables. Users looking to replicate should have experience in Python and R Programming and can refer to the GitHub link in the Supplementary Material for replication.

## Conclusion

We developed a claims-based classifier to identify stroke severity in AIS patients using discharge mRS. Importantly, we partially addressed potential bias by accounting for hospital-level patterns in sampling using mRS completion rates. Our classifier has expanded on previous research by using PCNASP and Medicare-linked data from several states to assess stroke severity.

## Data Availability

Restrictions apply to the availability of these data, which were used under license for this study. Medicare data are available through CMS with their permission. PCNASP data are available through the CDC with their permission.

## ACKNOWLEDGEMENTS FUNDING

This study was supported by the NIH (1R01AG073410-01)

## Supplemental Materials

Link to GitHub Code to replicability Figures S1-S4

Tables S1-9

